# Predicting and elucidating the etiology of fatty liver disease using a machine learning-based approach: an IMI DIRECT study

**DOI:** 10.1101/2020.02.10.20021147

**Authors:** Naeimeh Atabaki-Pasdar, Mattias Ohlsson, Ana Viñuela, Francesca Frau, Hugo Pomares-Millan, Mark Haid, Angus G Jones, E Louise Thomas, Robert W Koivula, Azra Kurbasic, Pascal M Mutie, Hugo Fitipaldi, Juan Fernandez, Adem Y Dawed, Giuseppe N Giordano, Ian M Forgie, Timothy J McDonald, Femke Rutters, Henna Cederberg, Elizaveta Chabanova, Matilda Dale, Federico De Masi, Cecilia Engel Thomas, Kristine H Allin, Tue H Hansen, Alison Heggie, Mun-Gwan Hong, Petra JM Elders, Gwen Kennedy, Tarja Kokkola, Helle Krogh Pedersen, Anubha Mahajan, Donna McEvoy, Francois Pattou, Violeta Raverdy, Ragna S Häussler, Sapna Sharma, Henrik S Thomsen, Jagadish Vangipurapu, Henrik Vestergaard, Leen M ‘t Hart, Jerzy Adamski, Petra B Musholt, Soren Brage, Søren Brunak, Emmanouil Dermitzakis, Gary Frost, Torben Hansen, Markku Laakso, Oluf Pedersen, Martin Ridderstråle, Hartmut Ruetten, Andrew T Hattersley, Mark Walker, Joline WJ Beulens, Andrea Mari, Jochen M Schwenk, Ramneek Gupta, Mark I McCarthy, Ewan R Pearson, Jimmy D Bell, Imre Pavo, Paul W Franks

## Abstract

**Background:** Non-alcoholic fatty liver disease (NAFLD) is highly prevalent and causes serious health complications in type 2 diabetes (T2D) and beyond. Early diagnosis of NAFLD is important, as this can help prevent irreversible damage to the liver and ultimately hepatocellular carcinomas.

**Methods and Findings:** Utilizing the baseline data from the IMI DIRECT participants (n=1514) we sought to expand etiological understanding and develop a diagnostic tool for NAFLD using machine learning. Multi-omic (genetic, transcriptomic, proteomic, and metabolomic) and clinical (liver enzymes and other serological biomarkers, anthropometry, and measures of beta-cell function, insulin sensitivity, and lifestyle) data comprised the key input variables. The models were trained on MRI image-derived liver fat content (<5% or ≥5%). We applied LASSO (least absolute shrinkage and selection operator) to select features from the different layers of omics data and Random Forest analysis to develop the models. The prediction models included clinical and omics variables separately or in combination. A model including all omics and clinical variables yielded a cross-validated receiver operator characteristic area under the curve (ROCAUC) of 0.84 (95% confidence interval (CI)=0.82, 0.86), which compared with a ROCAUC of 0.82 (95% CI=0.81, 0.83) for a model including nine clinically-accessible variables. The IMI DIRECT prediction models out-performed existing non-invasive NAFLD prediction tools.

**Conclusions:** We have developed clinically useful liver fat prediction models (see: www.predictliverfat.org) and identified biological features that appear to affect liver fat accumulation.

## INTRODUCTION

Non-alcoholic fatty liver disease (NAFLD) is characterized by the accumulation of fat in hepatocytes in the absence of excessive alcohol consumption. NAFLD is a spectrum of liver diseases with its first stage, known as ‘simple steatosis’, defined as liver fat content >5% of total liver weight. Simple steatosis can progress to non-alcoholic steatohepatitis (NASH), fibrosis, cirrhosis, and eventually hepatocellular carcinoma. In NAFLD, triglycerides accumulate in hepatocytes and liver insulin sensitivity is diminished, promoting hepatic gluconeogenesis, thereby raising the risk of type 2 diabetes (T2D) or exacerbating the disease pathology in those with diabetes (1-5).

The prevalence of NAFLD is thought to be around 20-40% in the general population in Western countries, with an increasing trend across the world, imposing a substantial economic and public health burden (6-9). However, the exact prevalence of NAFLD has not been clarified, in part because liver fat is difficult to accurately assess. Liver biopsy, magnetic resonance imaging (MRI), ultrasounds and liver enzyme tests are often used for NAFLD diagnosis, but the invasive nature of biopsies, the high costs of MRI scans, the non-quantitative nature and low sensitivity of conventional ultrasounds, and the low accuracy of liver enzyme tests are significant limitations (10-12). To address this gap, several liver fat prediction indices have been developed, but none of these has sufficiently high predictive ability to be considered a gold standard (10).

The purpose of this study was to use machine learning to identify novel molecular features associated with NAFLD and combine these with conventional clinical variables to predict NAFLD. These models include those variables that are likely to be informative of disease etiology, some of which may be of use in clinical practice.

## METHODS AND MATERALS

### Participants (IMI DIRECT)

The primary data utilized in this study were generated within the IMI DIRECT consortium, which includes a multicenter prospective cohort study of 3029 adults recently diagnosed with T2D (n=795) or at high risk of developing the disease (n=2234). All participants provided informed consent and the study protocol was approved by the regional research ethics committees for each clinical study center. Details of the study design and the core characteristics are explained elsewhere (13, 14).

### Measures (IMI DIRECT)

A T2*-based multiecho technique was used to derive liver fat content from MRI (15, 16) and the percentage values were categorized into fatty (≥5%) or non-fatty concentrations (<5%) to define the outcome variable. We elected not to attempt quantitative prediction of liver fat content, as this would require a much larger dataset to be adequately powered. A frequently-sampled 75g oral glucose tolerance test (OGTT) or a frequently sampled mixed-meal tolerance test (MMTT) was performed, from which measures of glucose and insulin dynamics were calculated, as previously described (13, 14, 17). Liver fat data were available for 1514 IMI DIRECT participants (503 diabetic and 1011 non-diabetic). The distribution of the liver fat data among different centers and cohorts is shown in S1 Fig and S2 Fig. The list of the clinical input (predictor) variables (n=58), including anthropometric, plasma biomarkers and lifestyle factors, are shown in S1 Table. These clinical variables were controlled for center effect by deriving residuals from a linear model including each clinical variable per model; these residuals were then inverse normalized and used in subsequent analyses. A detailed overview of participant characteristics for the key variables is shown in Table 1 for all IMI DIRECT participants with MRI data. There were no substantial differences in characteristics between these participants and those from IMI DIRECT who did not have MRI data (see S2 Table).

**Table 1.** Characteristics of IMI DIRECT participants in the non-diabetes, diabetes and combined cohorts separated for fatty vs. non-fatty individuals. Values are median (interquartile range) unless otherwise specified.

| Characteristics | Non-diabetic cohort |  | Diabetic cohort |  | Combined cohorts |  |
| --- | --- | --- | --- | --- | --- | --- |
|  | Fatty | Non-fatty | Fatty | Non-fatty | Fatty | Non-fatty |
| N (%) | 344 (34) | 667 (66) | 296 (59) | 207 (41) | 640 (42) | 874 (58) |
| Age (yr) | 61 (56, 66) | 62 (56, 66) | 62 (55, 67) | 63 (58, 69) | 61 (56, 66) | 62 (56, 67) |
| Sex, *n* (% female) | 62 (18) | 134 (20) | 130 (44) | 86 (42) | 192 (30) | 220 (25) |
| Weight (kg) | 90.75<br>(81.50, 100.25) | 81.40<br>(75.67, 89.60) | 92.85<br>(81.47, 103.75) | 80.80<br>(73.00, 93.55) | 91.20<br>(81.50, 102.00) | 81.40<br>(74.03, 90.17) |
| Waist circ. (cm) | 105<br>(98, 112) | 97<br>(91, 103) | 107<br>(97, 115) | 97<br>(90, 107.25) | 106<br>(98, 113) | 97<br>(91, 103) |
| BMI (kg/m<sup>2</sup>) | 29.23<br>(26.91, 32.05) | 26.69<br>(24.75, 28.71) | 31.47<br>(28.37, 35.35) | 27.64<br>(25.53, 31.07) | 30.05<br>(27.53, 33.52) | 26.85<br>(24.91, 29.23) |
| SBP | 134.70<br>(125.3, 143.0) | 129.33<br>(120, 140) | 131<br>(122, 139.33) | 127.67<br>(117.67, 138.33) | 132.67<br>(124.00, 142.00) | 128.83<br>(119.33, 140.00) |
| DBP | 83.50<br>(79.33, 89.83) | 80.67<br>(75.67, 86.00) | 76.67<br>(72, 84) | 72.67<br>(67.17, 80.67) | 81.33<br>(5.33, 87.33) | 80.00<br>(73.33, 84.67) |
| HbA1c (mmol/mol) | 38 (36, 40) | 37 (35, 39) | 47 (44, 51) | 45 (42, 48) | 41 (37, 46) | 38 (36, 41) |
| Fasting glucose (mmol/L) | 5.90<br>(5.60, 6.30) | 5.70<br>(5.4, 6) | 7.20<br>(6.3, 7.9) | 6.70<br>(5.8, 7.6) | 6.30<br>(5.8, 7.2) | 5.80<br>(5.4, 6.3) |
| Fasting insulin (pmol/L) | 75.60<br>(54.30, 104.40) | 44.10<br>(27.75, 66.00) | 115.8<br>(75.8, 167.8) | 60.20<br>(40.85, 82.90) | 90.90<br>(61.2, 133.9) | 48.60<br>(30.00, 69.60) |
| 2hr glucose (mmol/L) | 6.55<br>(5.37, 8.20) | 5.70<br>(4.70, 6.80) | 9<br>(6.90, 10.65) | 7.90<br>(6.20, 9.90) | 7.40<br>(5.90, 9.60) | 6<br>(4.90, 7.50) |
| 2hr insulin (pmol/L) | 345.60<br>(198.40, 566.20) | 169.80<br>(100.2, 274.2) | 489.30<br>(297.40, 700.50) | 271<br>(166.40, 418.10) | 403.20<br>(236.60, 643.50) | 190.70<br>(110.80, 317.60) |
| Triglycerides (mmol/L) | 1.49<br>(1.13, 2.09) | 1.12<br>(0.86, 1.47) | 1.49<br>(1.01, 1.99) | 1.12<br>(0.86, 1.48) | 1.49<br>(1.08, 2.02) | 1.12<br>(0.86, 1.47) |
| ALT (U/L) | 21 (14, 29) | 15 (10, 20) | 25 (19, 33.25) | 20 (16, 24) | 23 (16, 32) | 16 (12, 22) |
| AST (U/L) | 29 (24, 37) | 25 (21, 29.75) | 24 (20, 30) | 22 (19, 27) | 26 (22, 33) | 24 (20, 29) |
| Alcohol intake, *n* ("never", "occasionally", "regularly") | 21, 68, 255 | 91, 133, 443 | 52, 81, 163 | 38, 45, 124 | 73, 149, 418 | 129, 178, 567 |
| Liver fat | 8.80 (6.60, 13) | 2.2 (1.50, 3.30) | 11.10 (7.30, 15.82) | 2.70 (1.95, 4) | 9.5 (6.80, 14.30) | 2.4 (1.60, 3.50) |

Genetic, transcriptomic, proteomic, and metabolomic datasets were used as input omic variables in the analyses. Buffy coat was separated from whole blood, and DNA was then extracted and genotyped using the Illumina HumanCore array (HCE24 v1.0); genotype imputation was performed using the Haplotype Reference Consortium (HRC) and 1000 Genome (1KG) reference panels. Details of the quality control (QC) steps for the genetic data are described elsewhere (14). Transcriptomic data were generated using RNA-sequencing from fasting whole blood. Only protein-coding genes were included in the analyses, as reads per kilobase of transcript per million mapped reads (RPKM). The targeted metabolomic data of fasting plasma samples were generated using the Biocrates Absolute*IDQ* p150 kit. Additionally, untargeted LC/MS-based metabolomics was used to cover a broader spectrum of metabolites. A combination of technologies and quantitative panels of protein assays were used to generate ‘targeted’ proteomic data. This included Olink’s proximity extension assays (18), sandwich immunoassay kits using Luminex technology (MerckMillipore and R&D Systems, Sweden), microfluidic ELISA assays (ProteinSimple, USA (19)), as well as protein analysis services from Myriad RBM (Myriad GmbH, Germany) and for hsCRP (MLM Medical Labs GmbH, Germany). In addition, protein data were generated by single-binder assays using highly multiplexed suspension bead arrays (20). This approach (denoted ‘exploratory’ proteomics) included a combination of antibodies targeting proteins selected by the consortium given published and unpublished evidence for association with glycemic-related traits. More information about data generation and QC of the transcriptomic, proteomic, and metabolomic data are described in the Supporting Information. Technical covariates for transcriptomics include *guanine-cytosine mean content, insert size, analysis lane* and *RNA integrity number, cell composition, date* and *center*. Technical covariates for proteomics were *center, assay, plate number* and *plate layout* (n=4), and for the targeted metabolites the technical covariates were *center* and *plate*. These technical covariates were used to correct the omics data and the residuals were then extracted from these models and inverse normalized prior to further analyses.

### Feature selection (IMI DIRECT)

We developed a series of NAFLD prediction models, comprised of variables that are available within clinical settings, as well as those not currently available in most clinics (see S3 Table). We had two strategies for selecting the clinical variables: i) we selected variables based on the clinical-accessibility and their established association with fatty liver from existing literature without applying statistical procedures for data reduction (models 1-3); ii) a pairwise Pearson correlation matrix was used for feature selection of the clinical variables by placing a pairwise correlation threshold of r>0.8. We then selected the variables we considered most accessible among those that were collinear (model 4). Feature selection was undertaken in the combined cohort (diabetic and non-diabetic) in order to maximize sample size and statistical power. Of 1514 participants with liver fat data, 1049 had all necessary clinical and multi-omics data for a complete case analysis. We used k-nearest neighbors (k-NN) (21) imputation method with k equal to 10 as a means to reduce the loss of sample size but found that this did not materially improve predictive power in subsequent analyses and determined not to include these imputed data. An overview of the pairwise correlations among the clinical variables available in 1049 IMI DIRECT Study participants is presented in Fig 1. The abbreviations used for the variables in the figure are defined in S1 Table.

**Fig 1.**
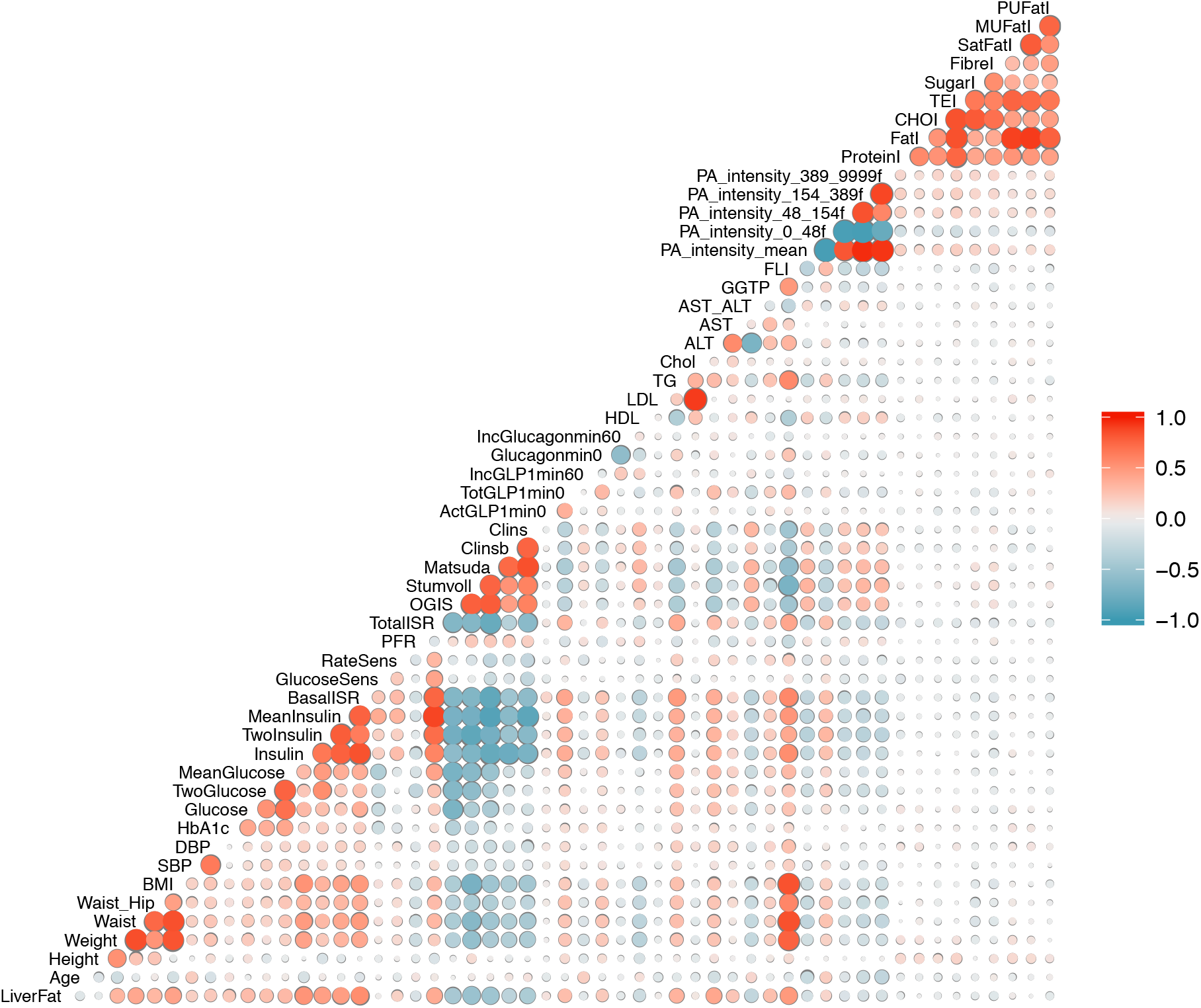
Pearson pairwise correlation matrix of clinical variables (data are inverse normal transformed) in the cohort combining participants with or without diabetes in IMI DIRECT (n=1049). The magnitude and direction of the correlation are reflected by the size (larger is stronger) and color (red is positive and blue is negative) of the circles respectively. The abbreviations used for the variables in the figure are defined in S1 Table.

The high-dimensionality nature of omics data also necessitated data reduction using the feature selection tool LASSO prior to building the model. LASSO is a regression analysis method that minimizes the sum of least squares in a linear regression model and shrinks selected beta coefficients (*β*_*j*_) using penalties (formula (1)). Minimizing the following value, LASSO excludes the least informative variables and selects those features of most importance for the outcome of interest (*y*) in a sample of n cases, each of which consists of m parameters. The penalty applied by *λ* can be any value from zero to positive infinity and is determined through a cross-validation step (22).

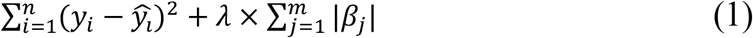

To minimize bias (for example by overfitting), we randomly divided the dataset and used 70% (n=735) for feature selection and 30% (n=314) for the model generation (see below). We selected these thresholds for partitioning the dataset in order to maximize the power to select the informative features. Stratified random sampling (23) based on the outcome variable was undertaken in order to preserve the distribution of the liver fat categories in the two feature selection and model generation sets. We selected LASSO, as a non-linear data reduction tool might lead to overfitting owing to the high dimensionality of omics data. LASSO was conducted with package glmnet in R (24) with a ten-fold cross-validation step for defining the *λ* parameter that results in the minimum value for the mean square error of the regression model.

Feature selection using LASSO was undertaken in each omics dataset (genetic, transcriptomic, proteomic and metabolomic) using 70% of the available data (models 5-18). For the genetic dataset, we first performed a genome-wide association study (GWAS) prior to LASSO in order to identify single-nucleotide polymorphisms (SNPs) tentatively associated with liver fat accumulation (*P*<5 × 10^78^). LASSO was then applied to these index variants for feature selection in 70% of the study sample. The individual SNP association analysis was conducted with rvTests v2.0.2 (25), which applies a linear mixed-model with an empirical kinship matrix to account for familial relatedness, cryptic relatedness, and population stratification. Only common variants with minor allele frequency (MAF) greater than 5% contributed to the kinship matrix. Liver fat data was log-transformed and then adjusted for age, age^2^, sex, center, body mass index (BMI) and alcohol consumption. These values were then inverse normal transformed and used in the GWAS analyses. S3 Fig and S4 Fig show the resulting Manhattan plot, depicting each SNP’s association with liver fat percentage and the quantile-quantile (QQ) plot of the GWAS results for liver fat. For the genetic data, 46 SNPs were selected out of the 623 SNPs with p-values < 5 × 10^78^. For the transcriptomics, 93 genes were selected out of 16,209 protein-coding genes. In the exploratory and targeted proteomics, 22 out of 377 and 48 out of 483 proteins were selected, respectively. In the targeted and untargeted metabolomic data, 39 out of 116 and 48 out of 172 were selected by LASSO, respectively.

### Model training and evaluation

The remaining 30% of the data was used to develop the binary prediction models for fatty liver (yes/no) with selected features used as input variables. We utilized the Random Forest supervised machine learning method, which is an aggregation of decision trees built from bootstrapped datasets (a process called ‘bagging’). Typically, two-thirds of the data are retained in these bootstrapped datasets and the remaining third is termed the ‘out of bag’ dataset (*OOB*), the latter of which is used to validate the performance of the model. To avoid over-fitting and improve generalizability, five-fold cross-validation was done for resampling the training samples and was repeated 5 times to create multiple versions of the folds. The number of trees was set to 1000 to provide an accurate and stable prediction. Receiver Operator Characteristic (ROC) curves were used to evaluate model performance by measuring the area under the curve (AUC). A ROC curve uses a combination of *sensitivity* (true positive rate) and *specificity* (true negative rate) to assess prediction performance. In our analysis, the Random Forest model is used to derive probability estimates for the presence of fatty liver. In order to make a class prediction, it is necessary to impose a cut-off above which fatty liver is deemed probable and below which it is considered improbable. The choice of cut-off influences both sensitivity and specificity for a given prediction model. We considered the effect of different cut-offs on these performance measurements. Additionally, we calculated the *F1 score* (26), which is the harmonic mean of precision (positive predictive value) and sensitivity, derived as follows:

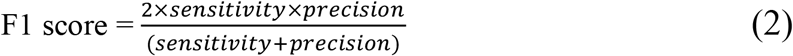

*Balanced Accuracy* was also evaluated, which is the proportion of individuals correctly classified (true positives and true negatives) within each class individually. Measurements of sensitivity, specificity, F1 score and balanced accuracy were computed and compared at different cut-offs for diabetic, non-diabetic and the combined cohorts. The variable importance was also determined via a “permutation accuracy importance” measure using Random Forest. In brief, for each tree, the prediction accuracy was calculated in the OOB test data. Each predictor variable was then permuted and the accuracy was recalculated. The difference in the accuracies was averaged over all the trees and then normalized by the standard error. Accordingly, a measure for variable importance is the difference in prediction accuracy before and after the permutation for each variable (27). Statistical analyses were undertaken using *R* software version 3.2.5 (28) and the Random Forest models were built using the Caret package (29). Fig 2 shows an overview of the different stages involved in the data processing and model training.

**Fig 2.**
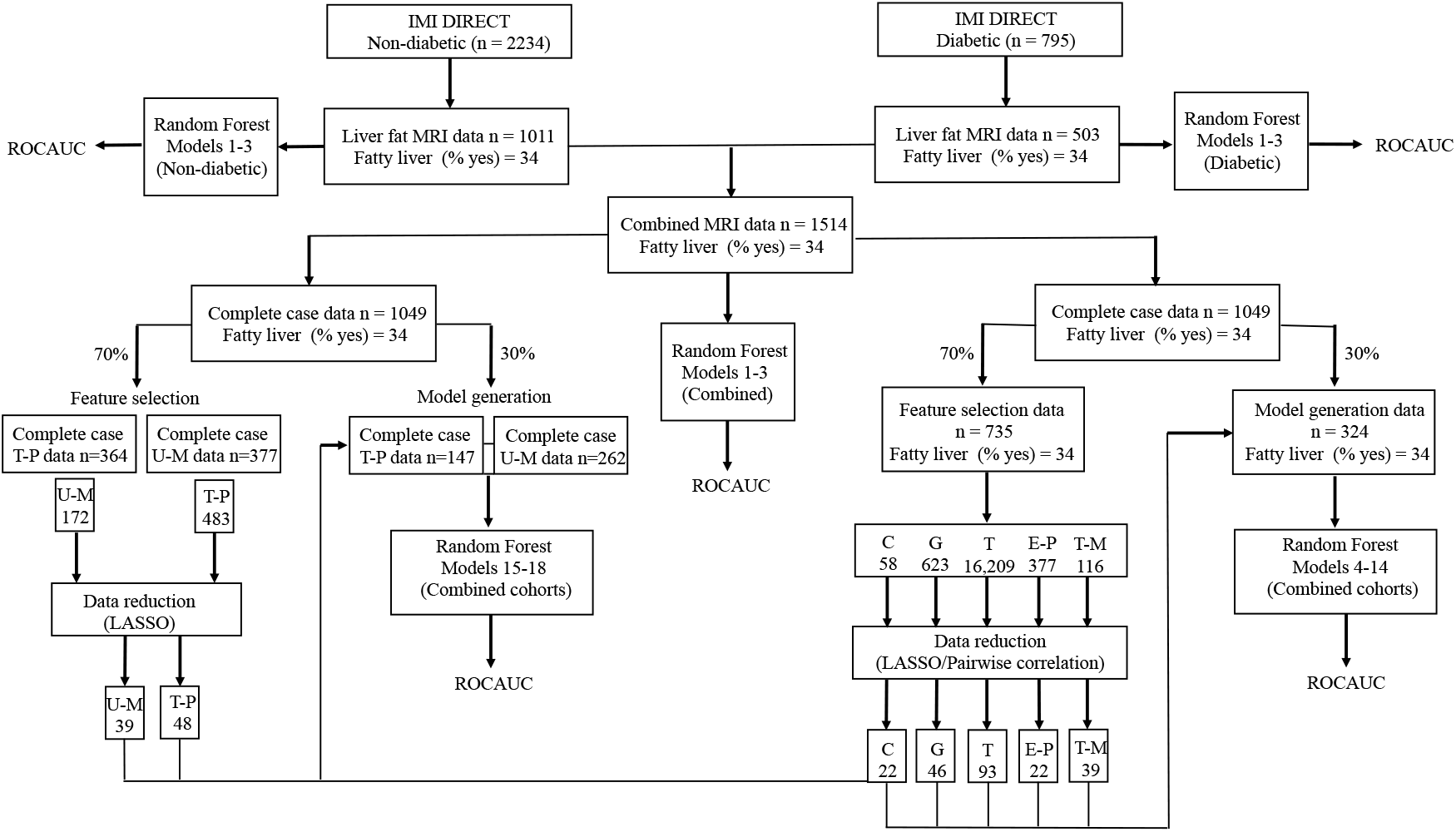
Overview of the different stages involved in data processing and model training: Clinical (C), Genetic (G), Transcriptomic (T), Exploratory Proteomic (E-P), Targeted Proteomic (T-P), Targeted Metabolomic (T-M) and Untargeted Metabolomic (U-M).

### Comparison with other fatty liver indices

Given the accessible data within the IMI DIRECT cohorts, several existing fatty liver indices could be calculated and compared with the IMI DIRECT prediction models. These included the *fatty liver index (FLI)* (30), *hepatic steatosis index (HSI)* (31) and *NAFLD liver fat score (NAFLD-LFS)* (32).

#### FLI

The FLI is commonly used to estimate the presence or absence of fatty liver (categorized into fatty (>=60) or non-fatty liver (<60) FLI units) (30). FLI uses data on plasma triglycerides (TG), waist circumference, BMI and serum gamma-glutamyl transpeptidase (GGTP) and is calculated as follows:

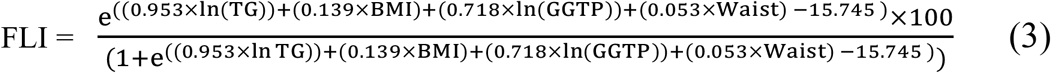

#### NAFLD-FLS

NAFLD-FLS was calculated using fasting serum (fs) insulin, aspartate transaminase (AST), alanine transaminase (ALT), T2D and metabolic syndrome (MS) (defined according to the International Diabetes Federation (33)) to provide an estimate of liver fat content. A NAFLD-FLS value above -0.64 is considered to indicate the presence of NAFLD:

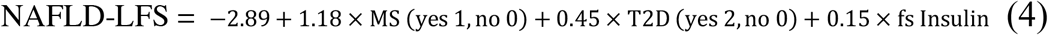

#### HSI

The HSI uses BMI, sex, T2D diagnosis (yes/no) and the ratio of ALT to AST and calculated as follows:

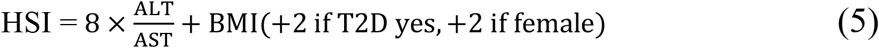

HSI values above 36 are deemed to indicate the presence of NAFLD.

### External validation (UK Biobank cohort)

The UK Biobank cohort (34) was used to validate the clinical prediction models (models 1 and 2) derived using IMI DIRECT data (UK Biobank application ID: 18274). The same protocol and procedure have been used to quantify MRI-derived liver fat in IMI DIRECT and UK Biobank (16). In addition, we validated the FLI and HSI using UK Biobank data. Field numbers for the UK Biobank variables used in the validation step can be found in the S4 Table. The data analysis procedures used for the UK Biobank validation analyses mirror those used in IMI DIRECT (as described above).

## RESULTS

The following section describes fatty liver prediction models that are likely to suit different scenarios. We focus on a basic model (model 1), which includes variables that are widely available in both clinical and research settings. Models 2 and 3 focus on variables that could in principle be accessed within the clinical context, but which are not routinely available in the clinical setting at this time. Model 4 includes clinical variables, more detailed measures of glucose and insulin dynamics, and physical activity. Models 5 to 18 are more advanced models that include omic predictor variables alone or in combination with clinical predictor variables. See S3 Table for a full description of models.

### Clinical models (Models 1-3)

We developed models 1-3 for NAFLD prediction, graded by perceived data accessibility for clinicians. These models were developed on the full dataset without applying any statistical procedures for feature selection. Model 1 includes six non-serological input variables: waist circumference, BMI, systolic blood pressure (SBP), diastolic blood pressure (DBP), alcohol consumption and diabetes status. Model 2 includes eight input variables: waist circumference, BMI, TG, ALT, AST, fasting glucose (or hemoglobin A1C (HbA1c) if fasting glucose is not available), alcohol consumption and diabetes status. Model 3 includes nine variables: waist circumference, BMI, TG, ALT, AST, fasting glucose, fasting insulin, alcohol consumption and diabetes status. The three clinical models along with FLI, HSI and NAFLD-LFS indices were applied to the non-diabetic and diabetic cohort datasets separately, as well as in the combined dataset; the ROCAUC results are presented in Fig 3. Model 1 yielded a ROCAUC of 0.73 (95% confidence interval (CI)=0.72, 0.75) in both cohorts combined. Adding serological variables to model 2 (with either fasting glucose or HbA1c) for the combined cohorts yielded the ROCAUC of 0.79 (95% CI=0.78, 0.80). Model 3 (fasting insulin added) yielded a ROCAUC of 0.82 (95% CI=0.81, 0.83) in the combined cohorts. The FLI, HSI and NAFLD-LFS had the ROCAUCs of 0.75 (95% CI=0.73, 0.78), 0.75 (95% CI=0.72, 0.77) and 0.79 (95% CI=0.76, 0.81) in the combined cohorts, respectively. The predictive performance of the clinical models 1-3, FLI, HSI and NAFLD-LFS in the non-diabetes and diabetes cohorts are presented in S5 Table.

**Fig 3.**
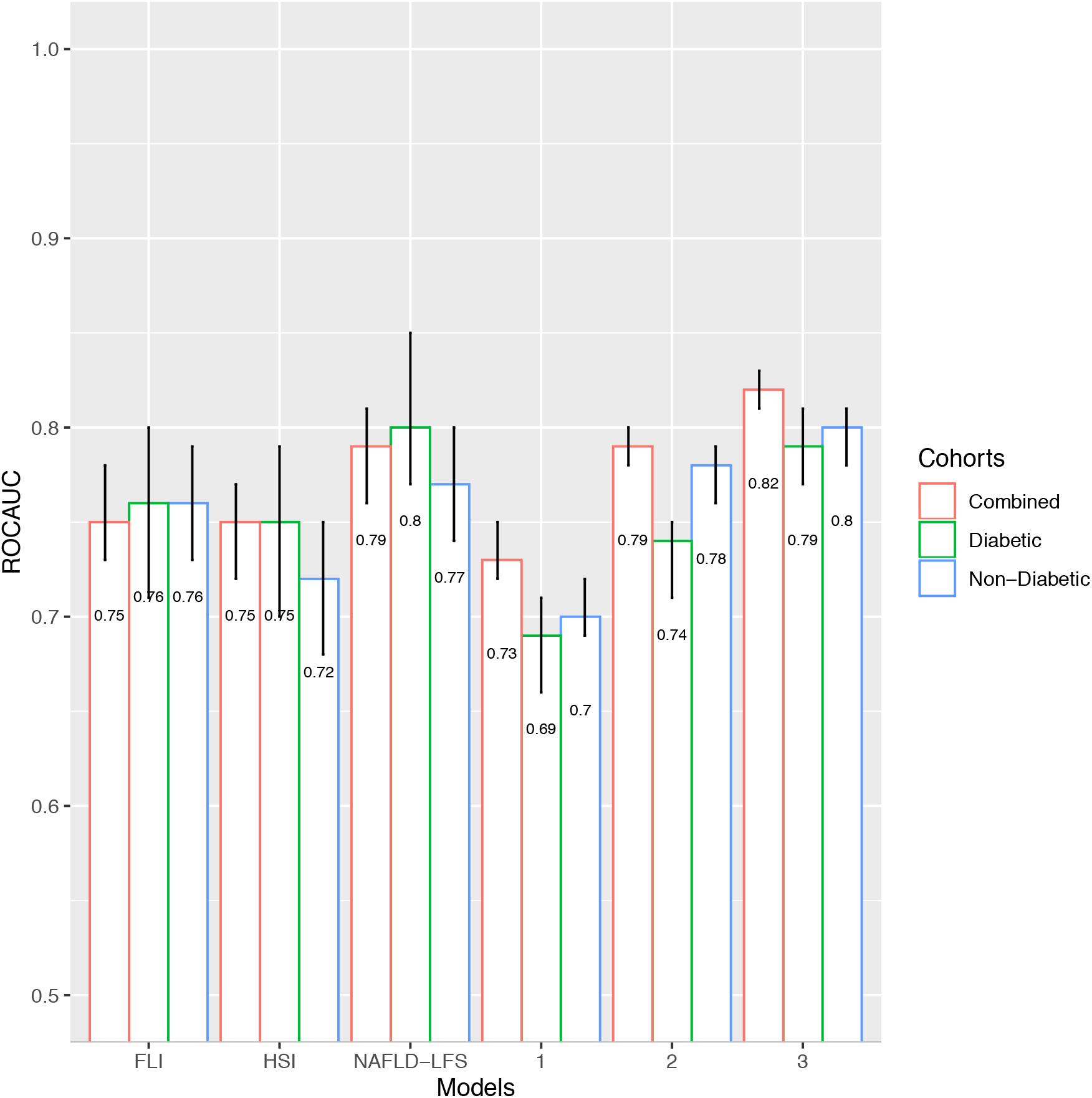
Receiver operator characteristic area under the curve (ROCAUC) with 95% confidence intervals (error bars) for the clinical models 1-3, FLI, HSI and NAFLD-LFS (x-axis) in the IMI DIRECT cohorts. Model 1 includes six non-serological input variables: waist circumference, BMI, SBP, DBP, alcohol consumption and diabetes status. Model 2 includes eight input variables: waist circumference, BMI, TG, ALT, AST, fasting glucose (or hemoglobin A1C (HbA1c) if fasting glucose is not available), alcohol consumption and diabetes status. Model 3 includes nine variables: waist circumference, BMI, TG, ALT, AST, fasting glucose, fasting insulin, alcohol consumption and diabetes status.

### Performance metrics

We further investigated sensitivity, specificity, balanced accuracy and F1 score (a score considering sensitivity and precision combined) metrics. These measurements were calculated for different cut-offs applied to the output of the Random Forest model (0.1, 0.2, 0.3, 0.4, 0.5, 0.6, 0.7, 0.8 and 0.9) using the clinical models (models 1-3) in the diabetic, non-diabetic and the combined cohorts. The performance metrics for models 1 and 2 are presented in S5 Fig and S6 Fig and for model 3 the metrics are presented in Fig 4. We aimed to find the optimal cut-off for these models based on the cross-validated balanced accuracy. The highest balanced accuracy for models 1-3 in the non-diabetic, diabetic and combined cohorts were observed at cut-offs of 0.4, 0.6 and 0.4, respectively (see Table 2).

**Table 2:**
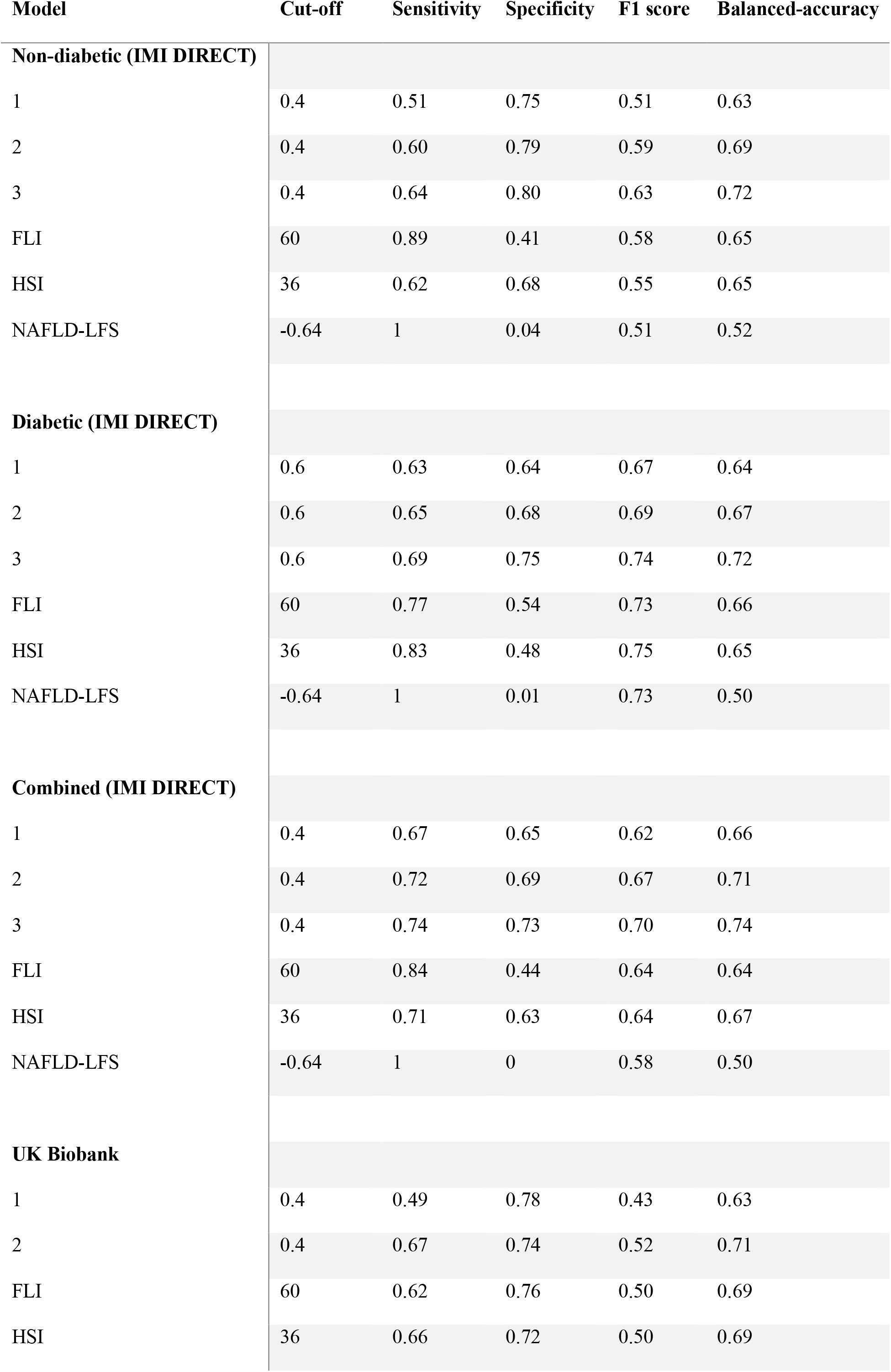
An overview of the prediction models’ performance metrics for all of the fatty liver indices in the IMI DIRECT and UK Biobank datasets

| Model | Cut-off | Sensitivity | Specificity | F1 score | Balanced-accuracy |
| --- | --- | --- | --- | --- | --- |
| **Non-diabetic (IMI DIRECT)** |  |  |  |  |  |
| 1 | 0.4 | 0.51 | 0.75 | 0.51 | 0.63 |
| 2 | 0.4 | 0.60 | 0.79 | 0.59 | 0.69 |
| 3 | 0.4 | 0.64 | 0.80 | 0.63 | 0.72 |
| FLI | 60 | 0.89 | 0.41 | 0.58 | 0.65 |
| HSI | 36 | 0.62 | 0.68 | 0.55 | 0.65 |
| NAFLD-LFS | -0.64 | 1 | 0.04 | 0.51 | 0.52 |
| **Diabetic (IMI DIRECT)** |  |  |  |  |  |
| 1 | 0.6 | 0.63 | 0.64 | 0.67 | 0.64 |
| 2 | 0.6 | 0.65 | 0.68 | 0.69 | 0.67 |
| 3 | 0.6 | 0.69 | 0.75 | 0.74 | 0.72 |
| FLI | 60 | 0.77 | 0.54 | 0.73 | 0.66 |
| HSI | 36 | 0.83 | 0.48 | 0.75 | 0.65 |
| NAFLD-LFS | -0.64 | 1 | 0.01 | 0.73 | 0.50 |
| **Combined (IMI DIRECT)** |  |  |  |  |  |
| 1 | 0.4 | 0.67 | 0.65 | 0.62 | 0.66 |
| 2 | 0.4 | 0.72 | 0.69 | 0.67 | 0.71 |
| 3 | 0.4 | 0.74 | 0.73 | 0.70 | 0.74 |
| FLI | 60 | 0.84 | 0.44 | 0.64 | 0.64 |
| HSI | 36 | 0.71 | 0.63 | 0.64 | 0.67 |
| NAFLD-LFS | -0.64 | 1 | 0 | 0.58 | 0.50 |
| **UK Biobank** |  |  |  |  |  |
| 1 | 0.4 | 0.49 | 0.78 | 0.43 | 0.63 |
| 2 | 0.4 | 0.67 | 0.74 | 0.52 | 0.71 |
| FLI | 60 | 0.62 | 0.76 | 0.50 | 0.69 |
| HSI | 36 | 0.66 | 0.72 | 0.50 | 0.69 |

**Fig 4.**
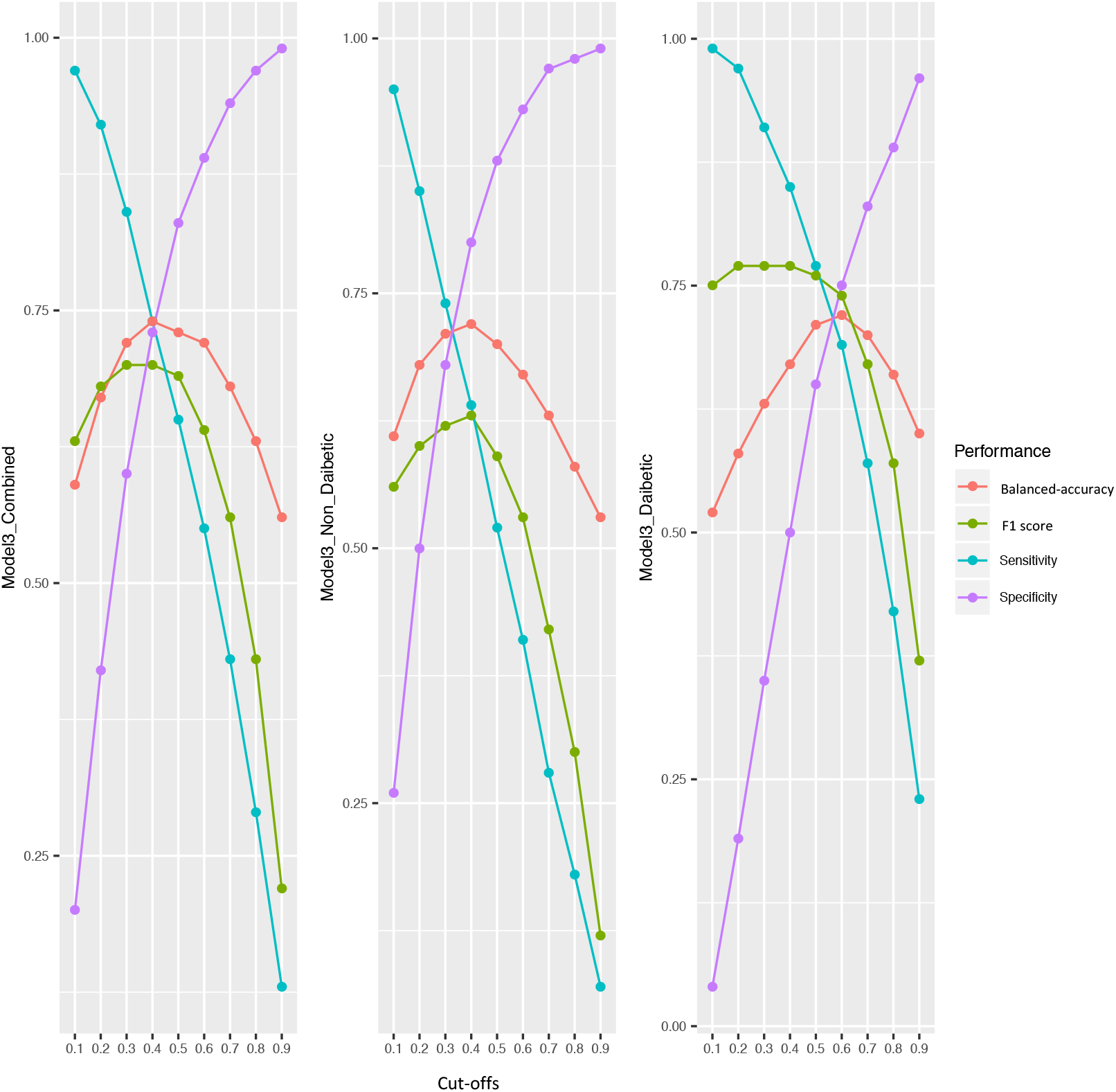
Measurements of F1, sensitivity, specificity and balanced accuracy (y-axis) at different cut-offs (x-axis) for model 3 in the diabetic, non-diabetic and the combined cohorts of the IMI-DIRECT. The measurements are calculated by defining the predicted probabilities of fatty liver equal or above these cut-offs as fatty and below as non-fatty.

Measurements of sensitivity, specificity, F1 score and balanced accuracy were computed for FLI, HSI and NAFLD-LFS indices and compared with those of the clinical models (1-3). These measurements were computed at the optimal cut-off values for these indices: -0.640 for NAFLD-LFS; 60 for FLI and 36 for HSI, respectively. A comprehensive overview of the prediction models’ performance metrics for all of the fatty liver indices listed above is shown in Table 2.

### Validation in UK Biobank and IMI DIRECT

Liver fat data were available in 4617 UK Biobank participants (1011 with >=5% and 3606 with <5% liver fat). Of these individuals, 4609 had all the required variables to replicate the clinical model 1. To perform model 2 either with fasting glucose or HbA1c, 3807 participants had data available for a complete case analysis. Given the limited availability of variables in the UK Biobank dataset, only models 1 and 2 of the NAFLD prediction models we developed could be externally validated. To facilitate this validation analysis, the Random Forest models developed in the IMI DIRECT cohorts were used to predict the liver fat category (fatty vs. non-fatty) for the UK Biobank participants. The performance of FLI and HSI was also tested in the UK Biobank cohort. We validated both models 1 and 2 in the UK Biobank cohort with a similar ROCAUC as seen in the IMI DIRECT dataset. The ROCAUCs were 0.71 (95% CI= 0.69, 0.73), 0.79 (95% CI= 0.77, 0.80), and 0.78 (95% CI= 0.76, 0.79), for model 1 and model 2 (with fasting glucose or with HbA1c), respectively. The FLI had a ROCAUC of 0.78 (95% CI= 0.76, 0.80), which is similar to model 2. The HSI yielded a ROCAUC of 0.76 (95% CI= 0.75, 0.78).

Measurements of sensitivity, specificity, F1 score and balanced accuracy were also computed at the optimal cut-off values for these models: 0.4 for clinical models 1and 2; 60 for FLI; 36 for HSI, respectively (see Table 2).

#### Omics models separately or in combination with clinical variables (models 5-14)

More advanced models using omics data were also developed. These models were generated using the omics features selected by LASSO in the combined cohorts. The models include only omics or include omics plus 22 clinical variables as the input variables. These clinical variables, selected based on the pairwise Pearson correlation matrix, are: BMI, waist circumference, SBP, DBP, alcohol consumption, ALT, AST, GGTP, HDL, TG, fasting glucose, 2-hour glucose, HbA1c, fasting insulin, 2-hour insulin, insulin secretion at the beginning of the carbohydrate challenge tests (OGTT or MMTT), insulin sensitivity 2-hour OGIS, mean insulin clearance during the OGTT/MTT,fasting glucagon concentration,fasting plasma total GLP-1 concentration,and mean physical activity intensity. Diabetes status(non-diabetic/diabetic) was also included as a clinical predictor in the models, given that analyses were undertaken in the combined diabetic and non-diabetic cohorts. The ROCAUCs for these models (models 4-14) are shown in Fig 5. The clinical model with the 22 selected clinical variables (model 4) yielded in ROCAUC of 0.79 (95% CI= 0.76, 0.81). Omics models with only the genetic (model 5), transcriptomic (model 7), proteomic (model 9) and targeted metabolomic (model 11) data as input variables resulted in ROCAUCs of 0.69 (95% CI= 0.66, 0.71), 0.72 (95% CI= 0.69, 0.74), 0.74 (95% CI= 0.71, 0.76) and 0.70 (95% CI= 0.67, 0.72), and respectively. Including all the omics variables in one model (model 13) resulted in a ROCAUC of 0.81 (95% CI= 0.76, 0.84). Adding the clinical variables to each omics model improved the prediction ability; models with the clinical variables plus genetic (model 6), transcriptomic (model 8), exploratory proteomic (model 10) and targeted metabolomic (model 12) resulted in ROCAUCs of 0.82 (95% CI= 0.80, 0.84), 0.81 (95% CI= 0.79, 0.83), 0.80 (95% CI= 0.78, 0.83) and 0.80 (95% CI= 0.77, 0.82), respectively. The highest performance was observed for model 14 (ROCAUC of 0.84, 95% CI= 0.82, 0.86). The variable importance for model 14 is presented in Fig 6, which shows that measures of insulin secretion rank amongst the highest of all input variables. Rankings for the individual clinical and omics variables are presented in S7-13 Figs.

**Fig 5.**
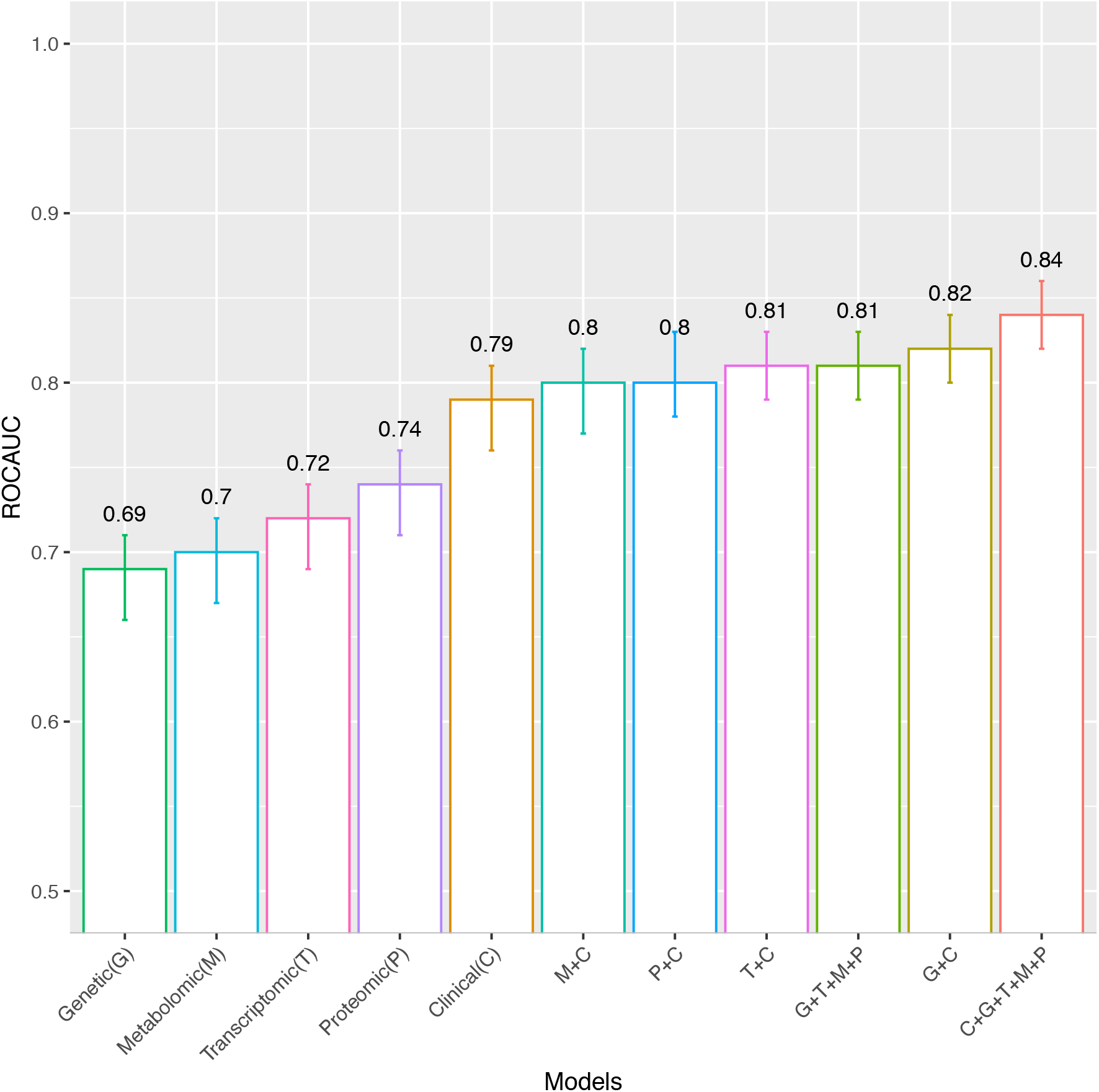
Receiver operator characteristic area under the curve (ROCAUC, y-axis) with 95% confidence intervals (error bars) for the Clinical model (C) with the 22 selected clinical variables (model 4), for the omics models separately (Genetic (G) (model 5), Transcriptomic (T) (model 7), Exploratory Proteomic (P) (model 9) and Targeted Metabolomic (M) (model 11)), for all omics together (G+T+M+P)(model 13) and for omics combined with the clinical model (C+G (model 6), C+T (model 8), C+P (model 10), C+M (model 12) and C+G+T+M+P (model 14)) in the cross-validated test data of IMI DIRECT combined cohorts.

**Fig 6.**
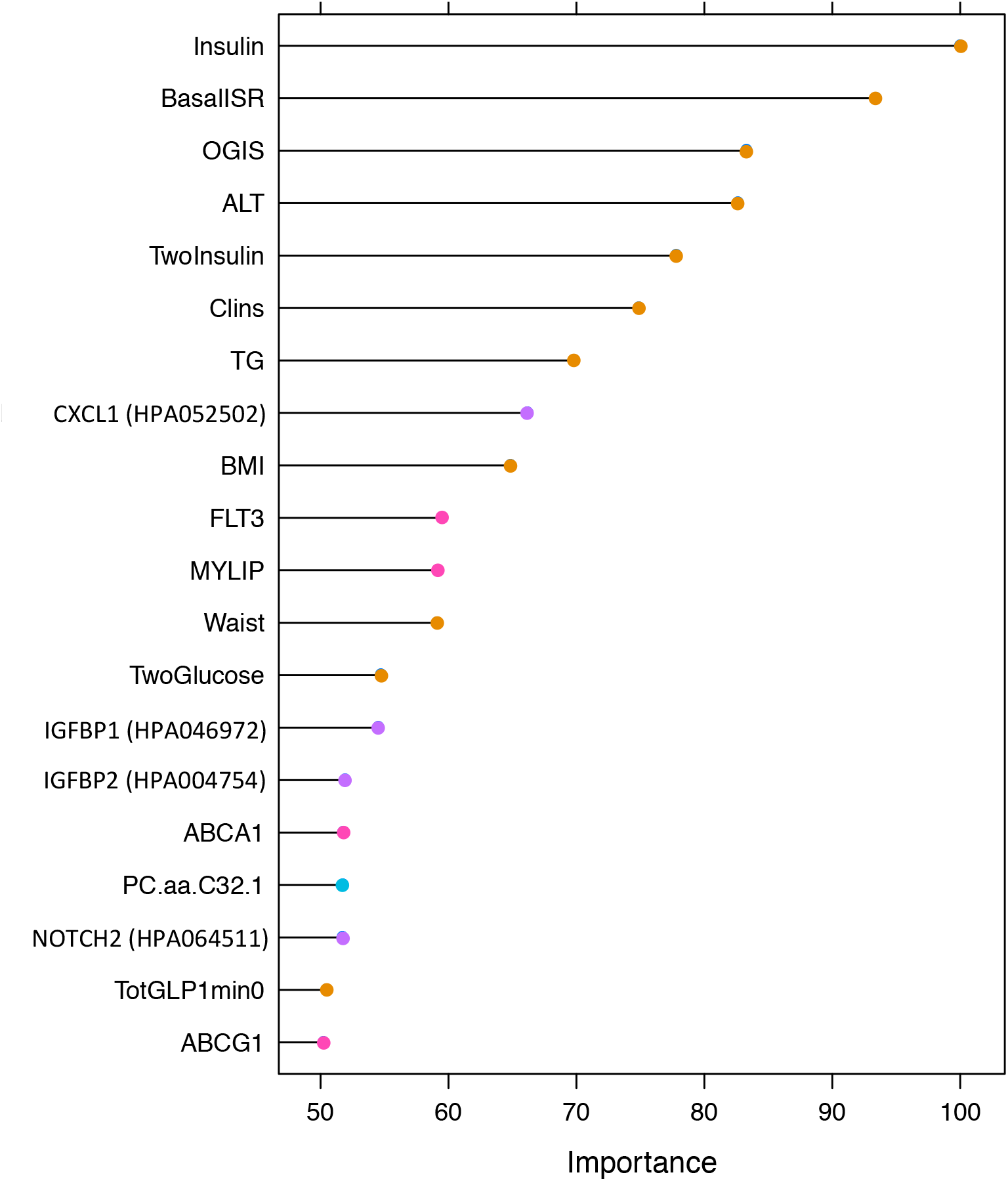
Variable importance for the Advanced model 14 with 222 omics and clinical input variables (clinical=22, genetic=46, transcriptomic=93, exploratory proteomic=22 and targeted metabolomic=39). The y-axis shows the top 20 predictors in the model. The x-axis shows the variable importance, calculated via a “permutation accuracy importance” measure using Random Forest as the difference in prediction accuracy before and after the permutation for each variable scaled by the standard error.

### Additional proteomic and metabolomic analyses (models 15-18)

Data from targeted proteomics and untargeted metabolomic data were further utilized to develop the omics models separately or in combination with the clinical data. However, as some participants lacked these omics data, their models were developed using a smaller data subset and were, hence, not included in the advanced (model 14) analyses. The complete case analysis was primarily defined on the availability of the 22 clinical variables (n=1049). Within this complete case set, 511 had a complete set of untargeted metabolomics data and 686 had a complete set of targeted proteomics data. Models with either targeted proteomic data only, or proteomic and clinical variables combined resulted in ROCAUCs of 0.81 (95% CI=0.78, 0.84) and 0.84 (95% CI=0.81, 0.87), respectively. The untargeted metabolomic model alone had a ROCAUC of 0.66 (95% CI=0.63, 0.69), which increased to 0.78 (95% CI= 0.75, 0.80) when the 22 clinical variables were added.

A web interface for the diagnosis of NAFLD was developed using the findings described above (www.predictliverfat.org), which we anticipate will render the models (1-3) developed here accessible for the wider community of clinicians and researchers.

## DISCUSSION

Using data from the IMI DIRECT consortium, we developed 18 diagnostic models for early-stage NAFLD. These models were developed to reflect different scenarios within which they might be used: these included both clinical and research settings, with the more complex (and less accessible) models having the greatest predictive ability. The models were successfully validated in the UK Biobank, where data permitted. Overall, the basic clinical variables proved to be stronger predictors of the fatty liver than more complex omics data, although adding omics data yielded the most powerful model, with very good cross-validated predictive ability (ROCAUC=0.84).

NAFLD is etiologically complex, rendering its prevention and treatment difficult, and diagnosis can require invasive and/or relatively expensive procedures. Thus, non-invasive and cost-effective prediction models with good sensitivity and specificity are much needed. This is especially important because if NAFLD is detected early, treatment through lifestyle interventions can be highly effective (35). However, simple NAFLD is usually asymptomatic and many patients only come to the attention of hepatologists when serious complications arise (36).

To date, several prediction models have been developed to facilitate the diagnosis of steatosis (thoroughly reviewed elsewhere (11)). FLI is the most well-established and commonly used index, initially developed using ultrasound-derived hepatic steatosis data (30). The FLI yielded similar predictive performance in the diabetic and non-diabetic cohorts of IMI DIRECT (ROCAUC ∼ 0.75).

Though commonly used for liver fat prediction, the FLI has a similar discriminative ability as waist circumference alone (37). Better discrimination can be obtained by incorporating additional serological and hemostatic measures, which is the case with the NAFLD-LFS (12), the SteatoTest (38) and the HSI (31), for example. Notwithstanding the added complexity and cost of these scores, the FLI, HSI and the NAFLD-LFS yielded similar predictive ability in a series of liver biopsy-diagnosed NAFLD cases (n=324) (32).

Omics technologies have been used in a small number of studies to identify molecular biomarkers of NAFLD (39-41). This includes tests utilizing genetic data such as FibroGENE for staging liver fibrosis (42), and tests using metabolomic data derived from liver tissue to differentiate simple hepatitis from NASH (43), as well as a multi-component NAFLD classifier using genomic, proteomic and phenomic data (41). Using data from IMI DIRECT, we explored the predictive ability of genetic, transcriptomic, proteomic and metabolomic data from the blood for the diagnosis of NAFLD. The top twenty features of each omic model are presented in the S7-13 Figs. The details of the LASSO selected features are summarized in the Supporting Information (Excel file). Reassuringly, several of the features that ranked highest have been previously described for their association with liver fat content or closely related traits; this includes *PNPLA3* gene variants (40, 44), fetal liver tyrosine kinase-3 (FLT3) transcripts (45), IGFBP1(46-48) and Lipoprotein lipase (Lpl) (49) proteins, and the metabolite glutamate (50). In the analysis of the targeted metabolites, phosphatidylcholines, including PC.aa.C32, PC.aa.C38, PC.aa.C40 and PC.aa.C42, glycerophospholipids and valine were amongst the highest-ranked metabolites that are known for their correlation with NAFLD and metabolic disorders (51, 52). For exploratory proteomics, the most important variables were proteins secreted into the blood, expressed by the liver as well as those leaking from the blood cells (53). The prediction model that only included targeted proteomic data (model 15) performed well (ROCAUC=0.81), rendering it an interesting candidate biomarker for future clinical tests. Among the top 20 most important proteins were many secreted into blood or leaked by the liver, as well as the pancreas, fat or muscle tissue (54).

The models developed here may be used for screening. In order to stratify people likely to have NAFLD who might then undergo more invasive and/or costly assessments, it would be necessary for the prediction model to have high sensitivity. However, the predictive utility of a given model can be further improved, by selecting model cut-offs that optimize sensitivity or specificity, as the two metrics rarely perform optimally at the same cut-off. This issue was apparent for models 1-3 in the current analyses, where we selected cut-offs that maximize balanced accuracy (considering both sensitivity and specificity); these features are especially important in screening algorithms, where the cost of false negatives can be high. Models 1-3 resulted in higher sensitivity in the diabetic cohort than the non-diabetic cohort, whereas the specificity was higher in the non-diabetic and in both cohorts combined than in the diabetic cohort.

It is noteworthy that the analytical methods deployed here required a complete case analysis, which diminishes sample size considerably and is, thus, a limitation of this approach; although imputing missing data here helped preserve sample size, it did not improve the prediction ability of the models, and we hence elected to use the complete case analysis. The linear Lasso method was used to minimize overfitting that can occur with high-dimensionality data, while Random Forests was used to identify non-linear associations where data structure allowed.

Heavy alcohol consumption is a key determinant of fatty liver but is unlikely to be a major etiological factor in IMI DIRECT owing to the demographics of this cohort. Nevertheless, a further limitation of this analysis is that alcohol intake was self-reported and may lack validity. To address this limitation, we removed all self-reported heavy alcohol consumers from the UK Biobank cohort and undertook sensitivity analyses, but this did not materially affect the results. A further consideration for future work is the impact lifestyle and medications are likely to have on the prediction of NAFLD. Here we considered lifestyle variables, but not medications. However, the use of medicines affecting liver fat is likely to be less in the non-diabetic than in the diabetic cohorts, yet the models fit better in the latter, suggesting that glucose-lowering medication use in the DIRECT cohorts does not have a major detrimental impact on prediction model performance.

In summary, we have developed prediction models for NAFLD that may have utility for clinical diagnoses and research investigations alike. Our finding that a model focused on proteomic data yielded high predictive utility may warrant further investigation. Our analysis also suggests that insulin sensitivity and beta-cell dysfunction may be involved in liver fat accumulation, which are at present not considered as features of conventional NAFLD risk models.

## Data Availability

Data cannot be shared publicly because of GDPR restrictions on data privacy.

## ACKNOWLEDGMENTS

We thank Mattias Borell for developing, logistical support and advice related with the web interface. We thank all the participants and study center staff in IMI DIRECT for their contribution to the study. We thank all the participants in the UK Biobank. This research was conducted using the UKBB Resource (application ID: 18274). For the proteomics analyses, we thank the entire staff of the Human Protein Atlas, the Plasma Profiling facility at Science for Life Laboratory and in particular Elin Birgersson, Annika Bendes, and Eni Andersson for technical assistance

## SUPPORTING INFORMATION CAPTIONS

**S1 Text**. QC of the transcriptomic, proteomic, and metabolomic

**S1 Fig**. Violin plot showing the distribution of liver fat percentage (y-axis) for the diabetic and non-diabetic cohorts (x-axis) of IMI DIRECT.

**S2 Fig**. Distribution of liver fat percentage (y-axis) among the different centers (x-axis) contributing to the IMI DIRECT cohorts.

**S3 Fig**. Manhattan plot showing SNPs associated with liver fat level (∼18 million imputed SNPs) in the IMI DIRECT cohorts. The chromosomal position is plotted on the x-axis and the statistical significance of association for each SNP is plotted on the y-axis. Red line indicates genome-wide significance level (5 × 10^−8^) and the blue line corresponds to the significance level of 5 × 10^−6^.

**S4 Fig**. Quantile-quantile (QQ) plot showing results of genome-wide association study (GWAS) for liver fat content in the IMI DIRECT consortium (1514 individuals). X-axis illustrates the expected distribution of *p-*values from association test across all the SNPs and the y-axis shows the observed *p*-values.

**S5 Fig**. Measurements of F1, sensitivity, specificity and balanced accuracy (y-axis) at different cut-offs (x-axis) for model 1 in the diabetic, non-diabetic and the combined cohorts of the IMI-DIRECT.

**S6 Fig**. Measurements of F1, sensitivity, specificity and balanced accuracy (y-axis) at different cut-offs (x-axis) for model 2 in the diabetic, non-diabetic and the combined cohorts of the IMI-DIRECT.

**S7 Fig**. Variable importance for the clinical model (only top 20)

**S8 Fig**. Variable importance for the genetic model (only top 20)

**S9 Fig**. Variable importance for the transcriptomic model (only top 20)

**S10 Fig**. Variable importance for the exploratory proteomic model (only top 20)

**S11 Fig**. Variable importance for the targeted metabolomic model (only top 20)

**S1 Table**. The list of the clinical input variables with the abbreviation used in the analyses and their meaning

**S2 Table**. Characteristics of the study in the non-diabetes, diabetes and combined cohorts separated for participants from IMI DIRECT who had MRI data vs. those who did not have MRI data. Values are median (interquartile range) unless otherwise specified.

**S3 Table**. Variables used to construct each of the NAFLD prediction models developed in the IMI DIRECT.

**S4 Table**. UK Biobank fields used in the analyses.

**S5 Table**. The ROCAUC results of the clinical models in the non-diabetes and diabetes cohorts separately.

**Supporting Information (Excel file)**. The details of the LASSO selected features of the omics layers.

## REFERENCES

1. Tilg H, Moschen AR. Insulin resistance, inflammation, and non-alcoholic fatty liver disease. Trends Endocrinol Metab. 2008;19(10):371–9.

2. Sattar N, Gill JM. Type 2 diabetes as a disease of ectopic fat? BMC Med. 2014;12:123.

3. Sattar N, Forrest E, Preiss D. Non-alcoholic fatty liver disease. BMJ. 2014;349:g4596.

4. Lucas C, Lucas G, Lucas N, Krzowska-Firych J, Tomasiewicz K. A systematic review of the present and future of non-alcoholic fatty liver disease. Clin Exp Hepatol. 2018;4(3):165–74.

5. Fazel Y, Koenig AB, Sayiner M, Goodman ZD, Younossi ZM. Epidemiology and natural history of non-alcoholic fatty liver disease. Metabolism. 2016;65(8):1017–25.

6. Bellentani S. The epidemiology of non-alcoholic fatty liver disease. Liver Int. 2017;37 Suppl 1:81–4.

7. Younossi ZM, Koenig AB, Abdelatif D, Fazel Y, Henry L, Wymer M. Global epidemiology of nonalcoholic fatty liver disease-Meta-analytic assessment of prevalence, incidence, and outcomes. Hepatology. 2016;64(1):73–84.

8. Younossi ZM. Non-alcoholic fatty liver disease - A global public health perspective. J Hepatol. 2019;70(3):531–44.

9. Younossi ZM, Tampi R, Priyadarshini M, Nader F, Younossi IM, Racila A. Burden of Illness and Economic Model for Patients With Nonalcoholic Steatohepatitis in the United States. Hepatology. 2019;69(2):564–72.

10. Castera L, Friedrich-Rust M, Loomba R. Noninvasive Assessment of Liver Disease in Patients With Nonalcoholic Fatty Liver Disease. Gastroenterology. 2019;156(5):1264–81 e4.

11. Castera L. Diagnosis of non-alcoholic fatty liver disease/non-alcoholic steatohepatitis: Non-invasive tests are enough. Liver Int. 2018;38 Suppl 1:67–70.

12. Kotronen A, Peltonen M, Hakkarainen A, Sevastianova K, Bergholm R, Johansson LM, et al. Prediction of non-alcoholic fatty liver disease and liver fat using metabolic and genetic factors. Gastroenterology. 2009;137(3):865–72.

13. Koivula RW, Heggie A, Barnett A, Cederberg H, Hansen TH, Koopman AD, et al. Discovery of biomarkers for glycaemic deterioration before and after the onset of type 2 diabetes: rationale and design of the epidemiological studies within the IMI DIRECT Consortium. Diabetologia. 2014;57(6):1132–42.

14. Koivula RW, Forgie IM, Kurbasic A, Vinuela A, Heggie A, Giordano GN, et al. Discovery of biomarkers for glycaemic deterioration before and after the onset of type 2 diabetes: descriptive characteristics of the epidemiological studies within the IMI DIRECT Consortium. Diabetologia. 2019;62(9):1601–15.

15. Thomas EL, Fitzpatrick JA, Malik SJ, Taylor-Robinson SD, Bell JD. Whole body fat: content and distribution. Prog Nucl Magn Reson Spectrosc. 2013;73:56–80.

16. Wilman HR, Kelly M, Garratt S, Matthews PM, Milanesi M, Herlihy A, et al. Characterisation of liver fat in the UK Biobank cohort. PLoS One. 2017;12(2):e0172921.

17. Robert W. Koivula IF, Azra Kurbasic, Ana Vinuela, Alison Heggie, Tue Hansen, Michelle Hudson, Anitra Koopman, Femke Rutters, Maritta Siloaho, Søren Brage, Adem Y. Dawed, Heather Ford, Giuseppe N. Giordano, Christopher J. Groves, Tarja Kokkola, Anubha Mahajan, Mandy H. Perry, Simone P. Rauh, Martin Ridderstråle, Harriet J. A. Teare, Louise Thomas, Andrea Tura, Henrik Vestergaard, Tom White, Jerzy Adamski, Jimmy Bell, Søren Brunak, Jacqueline Dekker, Emanouille Dermitzakis, Philippe Froguel, Gary Frost, Ramneek Gupta, Torben Hansen, Andrew Hattersley, Bernd Jablonka, Markku Laakso, Timothy J. McDonald, Oluf Pedersen, Andrea Mari, Mark I. McCarthy, Hartmut Ruetten, Imre Pavo, Mark Walker, Ewan Pearson, Paul W. Franks, for the IMI DIRECT consortium. Discovery of biomarkers for glycaemic deterioration before and after the onset of type 2 diabetes: an overview of the baseline data from the epidemiological studies within the IMI DIRECT Consortium. Diabetologia. 2018.

18. Assarsson E, Lundberg M, Holmquist G, Bjorkesten J, Thorsen SB, Ekman D, et al. Homogenous 96-plex PEA immunoassay exhibiting high sensitivity, specificity, and excellent scalability. PLoS One. 2014;9(4):e95192.

19. Aldo P, Marusov G, Svancara D, David J, Mor G. Simple Plex() : A Novel Multi-Analyte, Automated Microfluidic Immunoassay Platform for the Detection of Human and Mouse Cytokines and Chemokines. Am J Reprod Immunol. 2016;75(6):678–93.

20. Drobin K, Nilsson P, Schwenk JM. Highly multiplexed antibody suspension bead arrays for plasma protein profiling. Methods Mol Biol. 2013;1023:137–45.

21. Altman NS. An Introduction to Kernel and Nearest-Neighbor Nonparametric Regression. Am Stat. 1992;46(3):175–85.

22. Tibshirani R. Regression Shrinkage and Selection via the Lasso. Journal of the Royal Statistical Society. 1996;Vol. 58.

23. Setia MS. Methodology Series Module 5: Sampling Strategies. Indian J Dermatol. 2016;61(5):505–9.

24. Jerome Friedman TH, Robert Tibshirani. Regularization Paths for Generalized Linear Models via Coordinate Descent. Journal of Statistical Software, 33(1), 1–22. 2010.

25. Zhan X, Hu Y, Li B, Abecasis GR, Liu DJ. RVTESTS: an efficient and comprehensive tool for rare variant association analysis using sequence data. Bioinformatics. 2016;32(9):1423–6.

26. The statistical evaluation of medical tests for classification and prediction. [press release]. Oxford, UK: Oxford University Press 2003.

27. Strobl C, Boulesteix AL, Zeileis A, Hothorn T. Bias in random forest variable importance measures: illustrations, sources and a solution. BMC Bioinformatics. 2007;8:25.

28. Team RCR. A Language and Environment for Statistical Computing. 2013.

29. Max Kuhn. Contributions from Jed Wing SW, Andre Williams, Chris Keefer, Allan Engelhardt, Tony Cooper, Zachary Mayer, Brenton Kenkel, the R Core Team, Michael Benesty, Reynald Lescarbeau, Andrew Ziem, Luca Scrucca, Yuan Tang and Can Candan. caret: Classification and Regression Training. R package version 6.0-71. 2016.

30. Bedogni G, Bellentani S, Miglioli L, Masutti F, Passalacqua M, Castiglione A, et al. The Fatty Liver Index: a simple and accurate predictor of hepatic steatosis in the general population. BMC Gastroenterol. 2006;6:33.

31. Lee JH, Kim D, Kim HJ, Lee CH, Yang JI, Kim W, et al. Hepatic steatosis index: a simple screening tool reflecting nonalcoholic fatty liver disease. Dig Liver Dis. 2010;42(7):503–8.

32. Fedchuk L, Nascimbeni F, Pais R, Charlotte F, Housset C, Ratziu V, et al. Performance and limitations of steatosis biomarkers in patients with nonalcoholic fatty liver disease. Aliment Pharmacol Ther. 2014;40(10):1209–22.

33. Alberti KG, Zimmet P, Shaw J, Group IDFETFC. The metabolic syndrome--a new worldwide definition. Lancet. 2005;366(9491):1059–62.

34. Sudlow C, Gallacher J, Allen N, Beral V, Burton P, Danesh J, et al. UK biobank: an open access resource for identifying the causes of a wide range of complex diseases of middle and old age. PLoS Med. 2015;12(3):e1001779.

35. Lean ME, Leslie WS, Barnes AC, Brosnahan N, Thom G, McCombie L, et al. Primary care-led weight management for remission of type 2 diabetes (DiRECT): an open-label, cluster-randomised trial. Lancet. 2018;391(10120):541–51.

36. Araujo AR, Rosso N, Bedogni G, Tiribelli C, Bellentani S. Global epidemiology of non-alcoholic fatty liver disease/non-alcoholic steatohepatitis: What we need in the future. Liver Int. 2018;38 Suppl 1:47–51.

37. Motamed N, Sohrabi M, Ajdarkosh H, Hemmasi G, Maadi M, Sayeedian FS, et al. Fatty liver index vs waist circumference for predicting non-alcoholic fatty liver disease. World J Gastroenterol. 2016;22(10):3023–30.

38. Poynard T, Ratziu V, Naveau S, Thabut D, Charlotte F, Messous D, et al. The diagnostic value of biomarkers (SteatoTest) for the prediction of liver steatosis. Comp Hepatol. 2005;4:10.

39. Baranova A, Liotta L, Petricoin E, Younossi ZM. The role of genomics and proteomics: technologies in studying non-alcoholic fatty liver disease. Clin Liver Dis. 2007;11(1):209-20, xi.

40. Eslam M, Valenti L, Romeo S. Genetics and epigenetics of NAFLD and NASH: Clinical impact. J Hepatol. 2018;68(2):268–79.

41. Wood GC, Chu X, Argyropoulos G, Benotti P, Rolston D, Mirshahi T, et al. A multi-component classifier for nonalcoholic fatty liver disease (NAFLD) based on genomic, proteomic, and phenomic data domains. Sci Rep. 2017;7:43238.

42. Eslam M, Hashem AM, Romero-Gomez M, Berg T, Dore GJ, Mangia A, et al. FibroGENE: A gene-based model for staging liver fibrosis. J Hepatol. 2016;64(2):390–8.

43. Alonso C, Fernandez-Ramos D, Varela-Rey M, Martinez-Arranz I, Navasa N, Van Liempd SM, et al. Metabolomic Identification of Subtypes of Nonalcoholic Steatohepatitis. Gastroenterology. 2017;152(6):1449–61 e7.

44. Danford CJ, Yao ZM, Jiang ZG. Non-alcoholic fatty liver disease: a narrative review of genetics. J Biomed Res. 2018;32(5):389–400.

45. Al-Fayoumi S, Hashiguchi T, Shirakata Y, Mascarenhas J, Singer JW. Pilot study of the antifibrotic effects of the multikinase inhibitor pacritinib in a mouse model of liver fibrosis. J Exp Pharmacol. 2018;10:9–17.

46. Hagstrom H, Stal P, Hultcrantz R, Brismar K, Ansurudeen I. IGFBP-1 and IGF-I as markers for advanced fibrosis in NAFLD - a pilot study. Scand J Gastroenterol. 2017;52(12):1427–34.

47. Petaja EM, Zhou Y, Havana M, Hakkarainen A, Lundbom N, Ihalainen J, et al. Phosphorylated IGFBP-1 as a non-invasive predictor of liver fat in NAFLD. Sci Rep. 2016;6:24740.

48. Adamek A, Kasprzak A. Insulin-Like Growth Factor (IGF) System in Liver Diseases. Int J Mol Sci. 2018;19(5).

49. Chen Y, Huang H, Xu C, Yu C, Li Y. Long Non-Coding RNA Profiling in a Non-Alcoholic Fatty Liver Disease Rodent Model: New Insight into Pathogenesis. Int J Mol Sci. 2017;18(1).

50. Gaggini M, Carli F, Rosso C, Buzzigoli E, Marietti M, Della Latta V, et al. Altered amino acid concentrations in NAFLD: Impact of obesity and insulin resistance. Hepatology. 2018;67(1):145–58.

51. Imhasly S, Naegeli H, Baumann S, von Bergen M, Luch A, Jungnickel H, et al. Metabolomic biomarkers correlating with hepatic lipidosis in dairy cows. BMC Vet Res. 2014;10:122.

52. Koch M, Freitag-Wolf S, Schlesinger S, Borggrefe J, Hov JR, Jensen MK, et al. Serum metabolomic profiling highlights pathways associated with liver fat content in a general population sample. Eur J Clin Nutr. 2017;71(8):995–1001.

53. Uhlen M, Karlsson MJ, Hober A, Svensson AS, Scheffel J, Kotol D, et al. The human secretome. Sci Signal. 2019;12(609).

54. Uhlen M, Fagerberg L, Hallstrom BM, Lindskog C, Oksvold P, Mardinoglu A, et al. Proteomics. Tissue-based map of the human proteome. Science. 2015;347(6220):1260419.

